# Leveraging and partitioning polygenic risk scores to identify cancer-related proteins

**DOI:** 10.1101/2025.11.23.25340827

**Authors:** Diptavo Dutta, Jingning Zhang, Xinyu Guo, Mitchell Machiela, Kevin Brown, Josef Coresh, Alexis Battle, Elizabeth A. Platz, Nilanjan Chatterjee

**Affiliations:** Integrative Tumor Epidemiology Branch, Division of Cancer Epidemiology and Genetics, National Cancer Institute, USA; Department of Biostatistics, Johns Hopkins University, USA; Department of Quantitative and Computational Biology, University of Southern California, USA; Laboratory of Translational Genetics, Division of Cancer Epidemiology and Genetics, National Cancer Institute, USA; Department of Epidemiology, Johns Hopkins University, USA; Department of Oncology, Johns Hopkins University School of Medicine, and the Sidney Kimmel Comprehensive Cancer Center at Johns Hopkins, USA; Department of Biomedical Engineering, Johns Hopkins University, USA

## Abstract

**Background:** Large-scale genome-wide association studies (GWAS) have identified numerous common susceptibility variants associated with various cancers but underlying molecular mechanisms remain largely unknown.

**Methods:** Here we investigated the associations of susceptibility SNPs from 21 cancers with 4,955 plasma protein levels measured in cancer-free participants (N=8,664) from the Atherosclerosis Risk in Communities (ARIC) study. We used two complementary approaches, one based on analysis of associations of polygenic risk scores with the plasma proteome (pQTS) and the other based on a sparse canonical correlation analysis of the cancer-associated SNPs with the plasma proteome (ARCHIE), to detect potential mediating proteins and sub-networks.

**Results:** Across all cancers, we identified 90 associated proteins using pQTS of which 53 were distal (*trans*)-associations between cancer related SNPs and proteins. ARCHIE identified 19 significantly associated protein networks encompassing a broader set of 433 proteins often including the proteins identified by pQTS. We found that the proteins identified by pQTS and/or ARCHIE were enriched for relevant biological processes and cell types as well as cancer drivers and have somatic evidence of being associated with the respective cancers. For example, using SNPs associated with risk of basal cell carcinoma, we identified two protein sets having distinct functions: one primarily enriched in immune and inflammatory responses while the other enriched in pigmentation. Additionally, we identified proteins associated with multiple related cancers indicating potential pleiotropic protein activity.

**Conclusion:** Our analysis leverages known GWAS associations for cancers to identify protein networks underlying cancer risk and accordingly partition polygenic risk scores into mechanistic components. As detailed molecular data of relevant tissues, cell-types and developmental stage become increasingly available, similar approaches will prove to be important for identifying downstream molecular targets for GWAS variants and improve interpretation and research application of polygenic risk scores.

## Introduction

Large-scale genome-wide association studies (GWAS) have identified over a thousand common susceptibility variants across many different cancers and their subtypes^1,2^. A fundamental translational goal of GWAS is to pinpoint the potential causal genes and proteins that underlie these associations, thereby nominating potential therapeutic targets and informing prevention strategies^3^. However, this has proven to be particularly challenging given most GWAS-identified variants are non-coding^4–6^. Recent efforts to integrate data from intermediate molecular phenotypes, like gene expressions, protein levels, and metabolites, to understand the regulatory role of disease-related variants, has gained considerable traction in identifying molecular targets for diseases^7–10^. The emergence of data from large-scale “omics” resources, such as the Genotype-Tissue Expression (GTEx)^11^, INTERVAL^12^, UK Biobank^13^, the Atherosclerosis Risk in Communities (ARIC)^10,14^ study and others, in combination with current analytic tools, has facilitated expanded scrutiny of the effect of disease related variants on intermediate molecular phenotypes in shaping the genetic architecture of complex diseases, including cancers.

In particular, recent advancements in high-throughput technologies have allowed measurements on thousands of proteins in plasma samples collected from individuals participating in various large studies and biobanks^12,13,15^. This has ushered in new opportunities to investigate the role of proteins in complex diseases through complementary analytic approaches. Testing of direct associations between baseline measurements of proteins and incident events in prospective studies has identified many risk biomarkers across a host of different diseases and conditions, including cancers of different organ sites^16,17^. Further, a variety of genetic association studies have identified protein quantitative trait loci (pQTL), and the overlap of these pQTL with disease susceptibility loci have been exploited to examine the causal role of the proteins in disease etiology^18–20^. A recent study based on cis-pQTL of 2,094 circulating proteins and risk of nine cancers identified evidence of causality for 40 genes through a series of Mendelian randomization and co-localization analyses^21^.

To obtain insights on disease etiology, a complementary but less utilized approach for proteogenetic data has been to explore associations between polygenic risk scores (PRS) for diseases and the proteome for the identification of candidate proteins associated with disease risk, which we refer to here as protein quantitative trait score (pQTS)^22–25^. The approach can be particularly appealing for the discovery of important proteins under an “omnigenic” model where the effects of many genetic variants might converge on a much smaller set of disease-relevant ‘core’ genes^26,27^. Prior systematic pQTS analysis of cardiometabolic traits have identified 49 plasma proteins as potential mediators of genetic risk^23^, but such an approach has not been systematically utilized in studies of cancer. However, a limitation of the pQTS approach is that it may lack power when there are not just a few “core” proteins mediating most genetic effects. Instead, the effects may be distributed across heterogeneous subnetworks of proteins, each representing interconnected biological mechanisms that mediate the effects of only a small fraction of genetic loci. To this end, we recently showed that a powerful approach for identifying mediating genetic networks based on transcriptomic data is to explore *trans*-associations between disease-associated loci and gene expression levels based on sparse canonical correlation analysis^28^. The underlying methodology, ARCHIE, allows decomposing the overall PRS for a trait into multiple sub-PRS based on the patterns of association of the underlying variants with groups of molecular traits.

In this study, we leveraged proteogenetic data available from the Atherosclerosis Risk in Communities (ARIC) study^14^ to systematically investigate individual proteins and protein-networks that could mediate the effect of known risk loci of 21 different cancers. Across the analyzed cancers, we first identified target proteins by exploring associations between organ site-specific cancer PRS and 4,955 plasma proteins measured by the SomaScan® 5K assay. Further, for each cancer, we performed analysis using ARCHIE across the set of underlying susceptibility variants and the set of proteins to identify converging *trans*-effects of the SNPs through protein networks. Overall, the pQTS analysis identified a total of 63 proteins across 18 cancer types and the ARCHIE analysis identified a total of 433 protein networks across 14 cancer types. We carried out a series of follow-up analyses of these proteins to examine evidence of their overlaps with known cancer driver genes and identify potential pleiotropic effects of proteins across multiple cancers. We further conducted more detailed downstream analysis of two cancers (cervical cancer and basal cell carcinoma) to showcase how identified proteins can illuminate specific cancer-related pathways, cell types and processes.

## Results

### Overview of Methods

Our analysis is based on the plasma proteome data measured by SomaScan®5K assay from the 6,640 self-reported White and 2,024 for self-reported Black cancer-free participants with genetic data at visit 2 of ARIC study (See **Methods** for details and **Supplementary Table 1**). We refer to these participants as EA and AA respectively, with our primary analysis being focused on EA participants. The ARIC study was approved by the institutional review boards at each site and participants included in this analysis provided written informed consent.

We used a previously developed QC pipeline for preprocessing, log-transforming and adjustment of known and hidden confounder effects for the ARIC proteome data, resulting in 4,955 normalized plasma protein levels (See **Methods** for details)^10^. To identify target proteins associated with cancer-associated variants we employ two complementary methods as follows:

A. *pQTS (protein Quantitative Trait Score)*: For a given cancer (denoted by subscript *C*), we constructed PRS (r_c_) for each individual using the variants and weights provided by the PRS for cancer *C* in PGS catalog^29,30^ or otherwise. Then we use a simple linear model to identify the effect of the PRS on the protein level (p_k_) as:

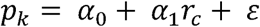 For each cancer analyzed, we use a Bonferroni correction threshold (1×10^−05^), correcting for 4,955 proteins, to identify significant pQTS associations (See **Methods**). Further, to evaluate whether a pQTS association is driven only by *trans*-associations between the variants constituting the PRS, and the proteins (*trans*-pQTS), we construct a restricted PRS r_c:trans_ using a subset of risk-associated PRS variants that are not within the cis-neighborhood (+/- 2.5 Mb) of a given protein k, and use a similar linear regression to evaluate its association as before:

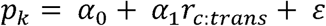 The same Bonferroni correction threshold is used to declare significant trans-pQTS.
B. *ARCHIE*: While the pQTS analysis can identify association of proteins proximal or distal to the SNPs in the PRS, we next focus on specifically identifying trans-associations of these SNPs and thus select potential downstream proteins. To identify proteins sets, *trans*-associated with subsets of the variants related to a given cancer, we use a sparse canonical correlation analysis (sCCA)^31–33^ based method, ARCHIE, that we have described in detail earlier^28^. For ARCHIE, we specifically focused on proteins that were distal (at least 2.5 Mb) from any SNP in the PRS of a given cancer and thus considered only *trans*-associations. Briefly, given a set of SNPs, constituting the PRS for a particular cancer, ARCHIE selects sparse linear combinations of SNPs (termed *SNP component*) which are strongly correlated with sparse linear combinations of proteins (termed *protein component*). We expect that the protein component selects downstream target proteins relevant to the cancer being analyzed, while the corresponding SNP component selects the variants that potentially regulate such proteins through intermediate mechanisms. Multiple pairs of such SNP and protein components (termed ARCHIE components together) can be extracted, each representing sets of proteins *trans*-associated with subsets of cancer related SNPs (See **Methods**). To evaluate which ARCHIE components represent cancer specific *trans*-association patterns, we employ a competitive testing framework, that tests for enrichment of correlation between the SNP and protein components against a polygenic background derived from GWAS significant variants, obtained through a resampling procedure (See **Methods**). This identifies the ARCHIE components that reflect *trans*-association patterns specific to the cancer being analyzed, potentially due to aggregation of multiple weaker *trans*-association between the corresponding SNPs and selected protein in the protein components.

### Identifying target proteins across 21 different cancers

In overall analysis with the EA participants, using variants significantly associated with 21 cancers, we identified 90 significant pQTS associations spanning 18 different cancers and 63 unique proteins (**Figure 1A**; **Supplementary Table 2**). Of these associations, majority (53; 58.9%) were *trans*-pQTS associations (**Figure 1B**), indicating that these proteins capture *trans*-effects for cancer-associated variants. Interestingly, for nearly 30% (16 of 53) associations, no individual variant was a strong *trans*-pQTL (p-value < 1×10^−06^), indicating that many of these proteins capture aggregated *trans*-effects of multiple risk variants. Since *trans*-associations can potentially identify downstream molecular targets, we further applied ARCHIE to the cancers that had at least one pQTS. Analysis with ARCHIE, identified significant components in 14 of the 18 cancers resulting in the selection of a broader set of 433 proteins overall, potentially *trans*-associated with subsets of cancer related SNPs constituting the corresponding PRS (**Supplementary Table 3**). Of these, a substantial fraction (38; 71.7%) overlap with the 53 proteins identified through *trans*-pQTS indicating high replicability of such identifications across methods.

**Figure 1:**
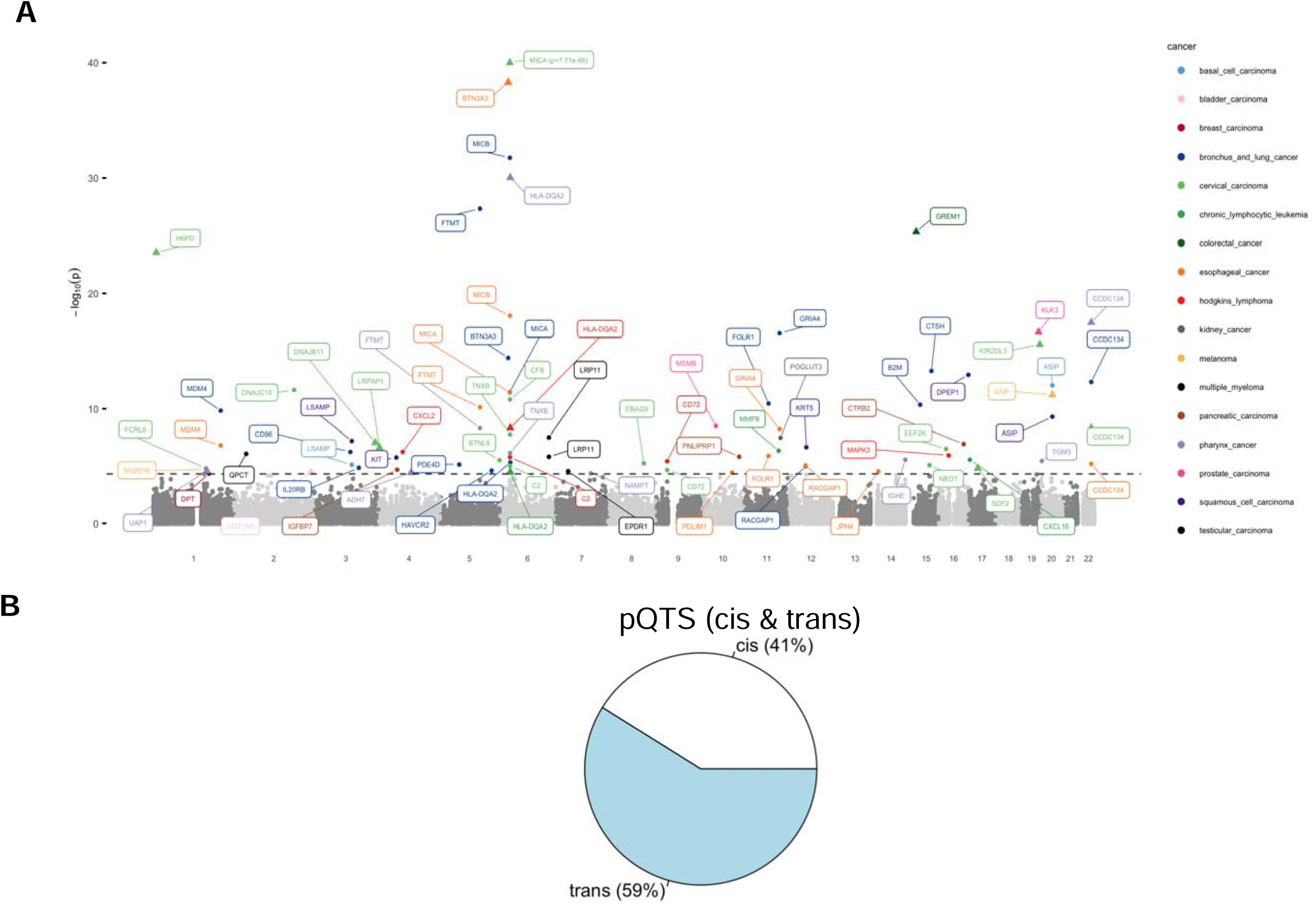
Proteins identified by pQTS across 21 different cancers. (A) Manhattan plot of the proteins identified by pQTS colored by the cancers. (B) Proportion of *cis* and *trans*-associations identified by pQTS

Existing databases, like COSMIC^34^, NCG^35^, have systematically catalogued key cancer driver and candidate driver genes by integrating evidence from somatic mutations, gene expression and cell lines. Across the proteins identified by either *trans*-pQTS or ARCHIE, we identify at least 83 (19.1%) established or candidate cancer driver genes (as reported in Network of Cancer Genes & Healthy Drivers), significantly higher than expected at random (p-value = 7.1×10^−03^; See **Methods**). Notable examples include *MDM4* (Mouse Double Minute 4), which plays a crucial role in the regulation of the tumor suppressor protein p53. Alterations in *MDM4*, including gene amplification, have been observed in various cancers and its overexpression is associated with several tumor progression and poor prognosis^36–38^. Among others, we also identify a proto-oncogene gene *KIT* (also known as *CD117*), which has a critical role in various cancer-related cellular processes, including cell survival, proliferation, and differentiation across multiple distinct cancers^39,40^.

### Pleiotropy

A systematic evaluation across cancers reveal that 47 proteins are identified in at least two cancers by either pQTS or ARCHIE, implying a potential pleiotropic role for such proteins. These included a higher proportion (18; 38.3%) of cancer drivers compared to overall associations. Among these was *CCDC134,* a protein that plays a role in the cell proliferation^41^ (**Figure 2B**) and was identified to be associated with 4 different cancers including pharynx cancer, bronchus and lung cancer, esophageal cancer and cervical carcinoma. This indicates a wide range of implications for the effect of the protein across multiple oncogenic mechanisms. However, we expect substantial overlap among proteins associated with similar cancers. For example, with three hematological cancers: chronic lymphocytic leukemia, multiple myeloma (MM) and Hodgkin’s lymphoma, we found 27 proteins to be associated with at least two of the three cancers and 11 proteins associated with all three cancers (**Figure 2A**), of which, two proteins *EEF2K* and *NRXN3* are candidate cancer driver genes^35^. Among the remaining, several proteins have been previously associated with different hematological and immune-related cancers and known immune response mechanisms like, *CXCL2* which is known to influence immune infiltration in tumors^42,43^; *IL21* has regulatory effects on immune cells and has been commonly associated with antitumor activity^44^. Data from Genomics of Therapeutics Response Portal (CTRP) database^45^ shows that IC50 for several antineoplastic drugs across 1001 cell lines, like panobinostat, dasatinib, PRIMA-1, afatinib, are highly correlated with the expressions of the 11 genes underlying the common proteins indicating potential for future drug repurposing studies.

**Figure 2:**
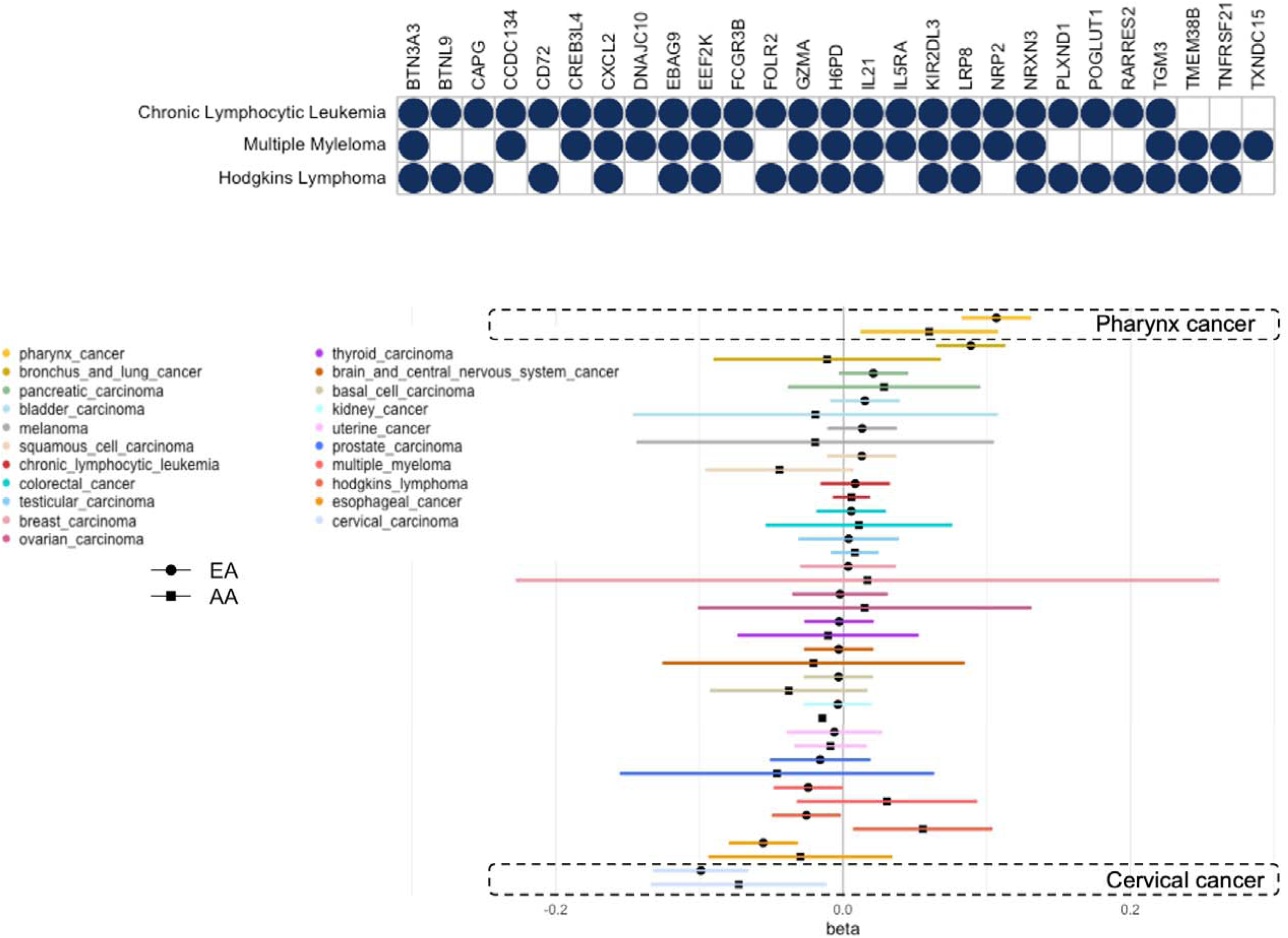
Pleiotropy and replication. (A) Plot showing 27 proteins (columns) identified in analysis with 3 hematological cancer (rows). Solid circles denote that protein was identified in either ARCHIE or pQTS analysis of corresponding cancer. (B) Forest plot showing the pQTS association of CCDC134 across cancers among self-reported White (EA) and Black (AA) participants. Cancers with significant associations in both are labeled.

### Replication in AA participants

We next investigated the replicability of the above results using data from AA participants in ARIC (N = 2,024) study although we anticipated fairly limited power of such analysis both due to substantially lower sample sizes and reduced transportability of PRS developed mainly from European ancestry population to African ancestry populations^46,47^. For this, we only investigated the replication of pQTS results in AA participants. Among the 90 significant pQTS associations identified in analysis with EA, we identified 22 (24.4%) proteins to have a nominal pQTS association (p-value < 0.05), while 64 (72%) had concordant direction of effects, in analysis with AA. Among the significant findings, was *GREM1,* which manifested significant (p-value < 1×10^−05^) pQTS association with colorectal cancer in both EA and AA, primarily driven by cis-variants. Another example is *CCDC134* which was identified to be associated with pharynx cancer and cervical carcinoma both in EA and AA (**Figure 2B**).

Next, we demonstrate as examples, results for two different cancers and the corresponding selected target proteins. Through several detailed downstream analyses, we show that the identified proteins have relevance to the overall genetic architecture of the respective cancers.

### Cervical cancer (CERC)

Cervical cancer (CERC) is the fourth most common cancer among women globally, with a majority of the cases being initiated by human papillomavirus (HPV) infection. Host genetics contributes a substantive proportion of CERC susceptibility, with a significant contribution from the biological processes related to immune response especially including genes in the HLA locus^48–50^. We used the PRS consisting of 10 SNPs^51^ with risk allele odds ratio ranging from 1.19 to 1.25 (mean = 1.22), all of them being located in the HLA region on chromosome 6. Using pQTS approach, we found that the PRS of CERC is associated with 19 different proteins at a Bonferroni correction threshold of 1×10^−05^ (**Figure 3A; Supplementary Table 2**) of which 6 proteins were in a cis-neighborhood of the HLA region. These included proteins like *MICA*, *CFB*, *TNXB* and *C2*, whose immunoregulatory functions have been extensively reported and can possibly contribute to the etiology of CERC^52,53^. The remaining 13 proteins were distal to any of the SNPs constituting the PRS, exhibiting trans-pQTS associations outside of the HLA region.

**Figure 3:**
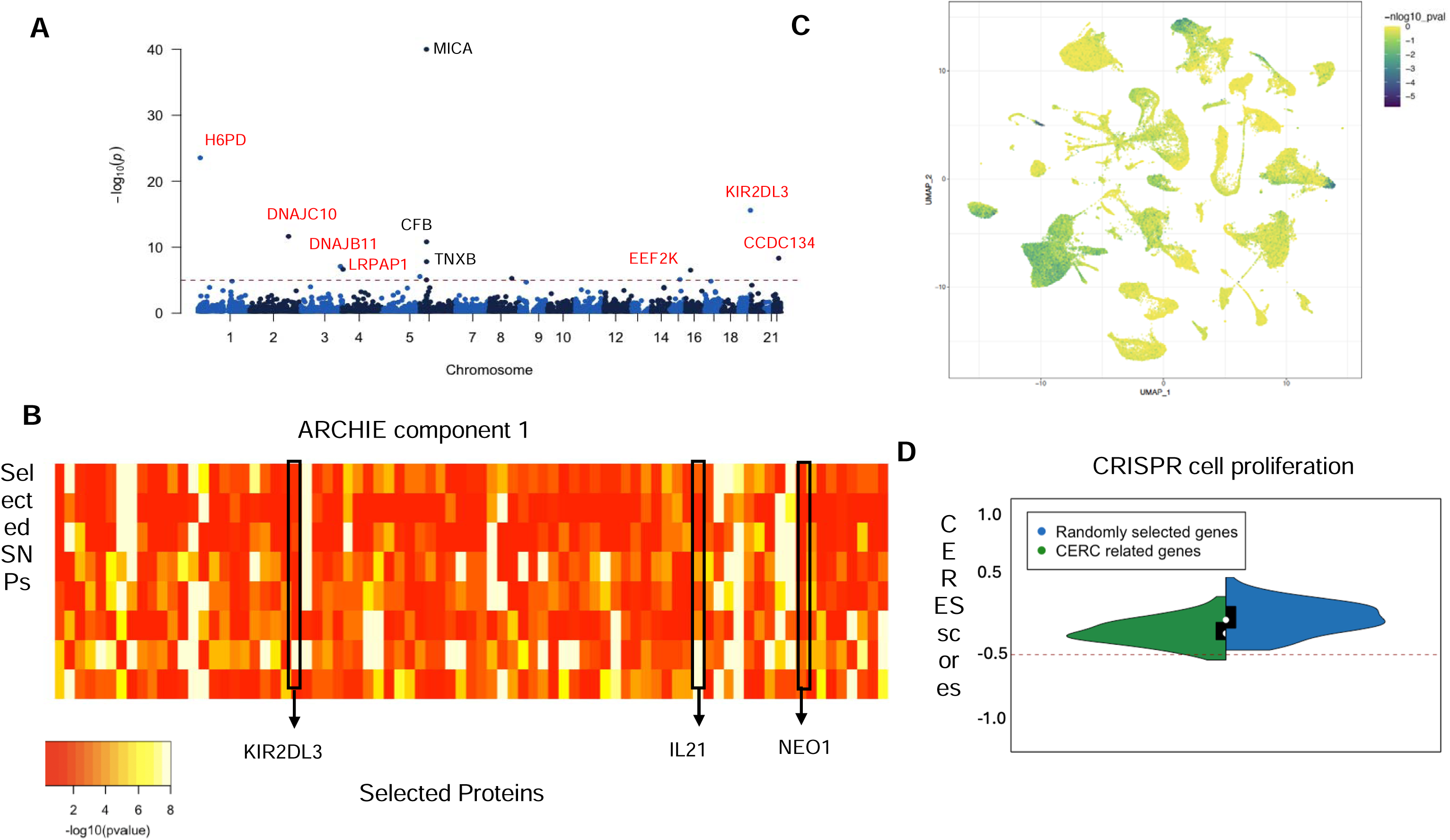
Proteins associated with Cervical Carcinoma (CERC) (A) Manhattan plot of the proteins identified by pQTS colored by significant cis (black) and trans (red)-associations. (B) −log10(trans-pQTL p-values) for proteins and SNPs selected by ARCHIE in the significant component. Proteins which are identified both by pQTS and ARCHIE, are highlighted. (C) scDRS analysis highlighting the cell populations whose expression are enriched for the significant proteins across pQTS and ARCHIE. (D) Distribution of CERES sc **Methods**) for genes selected by pQTS and ARCHIE (in green), in comparison to that for randomly selected genes (in blue).

Notable among the trans-pQTS associations is *KIR2DL3* (β = 0.14; p-value=2.6×10^−16^) a gene in the class of Killer Ig-like Receptors (KIR) which are known to be involved in regulating natural killer (NK) cells by interacting with HLA ligands. Genetic association studies, including that of cervical cancer, have previously suggested role of HLA-KIR interactions in etiology of various immune related diseases^54–56^. The current *trans*-pQTS analysis provides evidence for a proteomic level signature of the interaction between KIR and HLA ligands in etiology of CERC. Among the 13 *trans*-pQTS associations, 11 proteins had at least one strong trans-pQTL association (p-value < 1×10^−06^). The remaining 2 proteins, Neogenin 1 (*NEO1*) and Fc Receptor Like 6 (*FCRL6*), were identified possibly due to the convergence of signals from multiple weaker *trans*-associations, demonstrating the utility of pQTS approach, and have previously been linked to immune response, cancers, and cancer related processes^57–59^.

Analysis using ARCHIE identified one significant component, constituting 8 SNPs (out of 10 CERC-related SNPs) and 80 proteins (**Figure 3B**; **Supplementary Table 3**). All of the 13 distal proteins identified through trans-pQTS were identified as a part of the broader network of proteins selected by ARCHIE. However, the potential statistical advantage of ARCHIE over pQTS is that it can incorporate opposing effects of different SNPs on a given protein as well. For example, ARCHIE identifies *CREB3L4* that has strong opposing associations (*trans*-pQTL p-value < 1 × 10^−10^) with three different SNPs (positive for risk alleles of rs2523557 and rs1882; negative for risk allele of rs4713460). This protein is related to variety of immune functions and acts as a transcription factor targeting genes involved in angiogenesis and immune infiltration^60–62^, highlighting its potential role in CERC.

Follow up analysis including pathway enrichment (**Supplementary Table 4**) revealed that the 80 proteins identified by ARCHIE (including the 13 identified in *trans*-pQTS), are overrepresented in targets of key transcription factors like *CREB1* (FDR = 4.1×10^−02^) and *RELA* (FDR = 4.2×10^−02^) as well as several key cancer related pathways like cAMP signaling pathway (FDR = 3.2×10^−02^), TNF signaling pathway (FDR = 1.5×10^−02^), angiogenesis (FDR = 4.5×10^−02^), as well as immune response (FDR = 4.7×10^−06^). Analysis using scDRS^63^ revealed that the expressions of the 80 corresponding genes were significantly enriched in (**Supplementary Table 5**) single cell populations of T cells, dendritic cells, and macrophages (**Figure 3C**). Using data from cervical squamous cell carcinoma and endocervical adenocarcinoma in The Cancer Genome Atlas project^64^ (TCGA-CESC; N = 310), we found that the presence of somatic mutations in the set of 80 identified target proteins were nominally correlated with abundance of immune cells (**Supplementary Table 6**) like exhausted T cells (p-value = 2.4×10^−04^), Th1 cells (p-value = 2.5×10^−03^), and CD4 T cells (p-value = 1.7×10^−02^). Further, among the 80 target proteins identified, we found 13 to be candidate cancer driver genes (enrichment p-value= 4.8×10^−07^) indicating them to be associated with key cancer related mechanisms. Using CERES scores from knockout experiments on 13 CESC cell lines from DepMap, we found 7 genes showing evidence for essentiality (CERES < −0.5) of cell proliferation (**Supplementary Table 7**) and that the 80 target genes taken together, had a significantly lower CERES score (Wilcoxon p-value = 0.011; **Figure 3D**) compared to randomly selected genes, indicating that these genes have important effect on cell proliferation in CERC.

### Basal Cell Carcinoma (BCC)

Basal cell carcinoma (BCC) is the most common form of skin cancer, and an estimated 3.6 million cases are diagnosed each year in the U.S. Studies have revealed contributions of genetic factors, chronic immunosuppression, along with exposure to non-genetic risk factors have crucial role in BCC pathophysiology. We use the PRS consisting of 26 SNPs^65^ with risk allele odds ratio ranging from 1.57 to 1.76 (mean = 1.66). Using the pQTS approach, we found that the PRS of BCC is associated with two different proteins at a Bonferroni correction threshold of 1×10^−05^ (**Figure 4A; Supplementary Table 2**) of which *LSAMP* was identified through *trans*-pQTS and is an immunoglobulin that has been shown to act as tumor suppressor in several different cancers.

**Figure 4:**
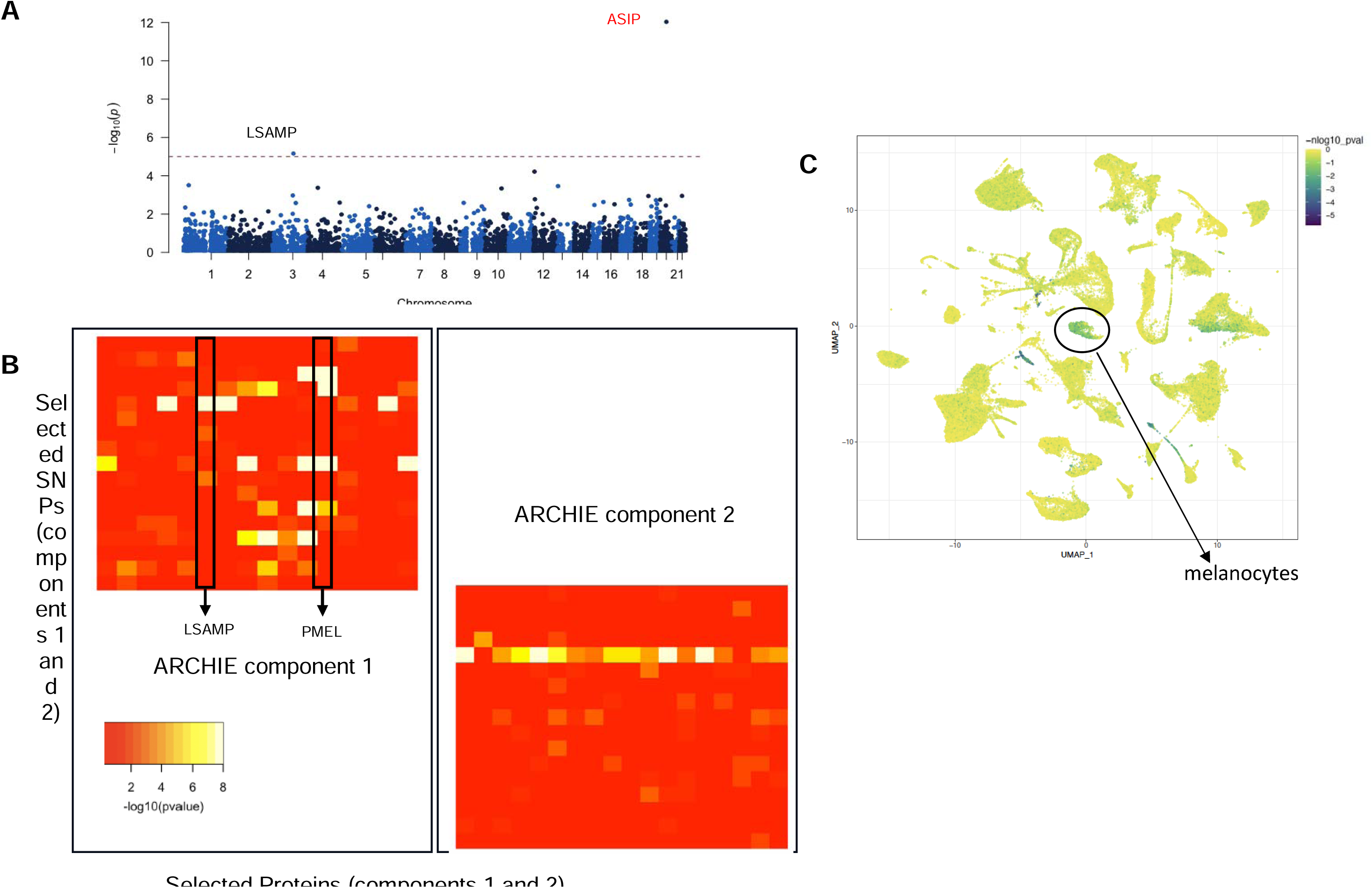
Proteins associated with Basal Cell Carcinoma (BCC) (A) Manhattan plot of the proteins identified by pQTS c lored by significant cis (black) and trans (red)-associations. (B) −log10(trans-pQTL p-values) for proteins and SNPs selected by ARCHIE in the significant component. (C) scDRS analysis highlighting the cells whose expression are enriched for the proteins selected by ARCHIE in component 1 with sub-population of melanocytes labelled.

ARCHIE identified two significant components, constituting 18 SNPs (out of 26 SCC-related SNPs) and 29 proteins overall which include *LSAMP* as a part of component 1, identified previously by *trans*-pQTS (**Figure 4B; Supplementary Table 3**). ARCHIE identifies several additional key proteins for BCC. For example, ARCHIE component 1 selects premelanosome (PMEL) protein which has strong (p-value < 1 × 10^−08^) opposing trans-pQTL associations with the risk allele of three different SNPs (positive for rs35407 and rs1126809; negative for risk alleles of rs4268748) and hence is not identified as a *trans*-pQTS. This protein is expressed primarily in pigment cells of the skin, crucial for pigmentation and has been well reported to be important for types of melanomas^66,67^.

Interestingly, we note that the proteins identified in the two ARCHIE components have markedly different functions. The proteins in ARCHIE component 1 are overrepresented for targets of *MITF* (FDR = 4.5×10^−06^) and genes involved in UV response (FDR = 5.9×10^−04^), indicating potential roles in pigmentation and processes involving skin cells in particular (**Supplementary Table 8**). This is also highlighted by the fact that scDRS reveals the expression of these genes to be enriched in single-cell populations of melanocytes among others (**Figure 4C**; **Supplementary Table 9**). In contrast, the proteins identified in ARCHIE component 2, which includes *IL21*, *CD8A*, and *IL36A* are primarily related to immune responses. The 15 proteins in ARCHIE component 2 are overrepresented in targets of several key cancer related and immune process related transcription factors like *E2F6* (FDR adjusted p-value = 3.74×10^−02^) and *CREB1* (FDR adjusted p-value = 2.5×10^−02^; **Supplementary Table 10**) and were differentially expressed in populations of T cells and macrophages. Given the marked separation in the biological processes and functions of the proteins in component 1 and 2, this provides a conceptual partition of the SNPs constituting the BCC PRS into broad pigmentation related processes, selected by component 1 and immune responses, selected by component 2.

Given the absence of publicly available large scale molecular profiling of BCC, we used data from Skin Cutaneous Melanoma in TCGA (TCGA-SKCM) to further evaluate the potential of the identified genes in skin cancer. However, we acknowledge that this might not be ideal for such comparison given notable etiological differences with BCC. We found somatic mutations in genes encoding the proteins selected by ARCHIE (including that by *trans*-pQTS as well), were significantly associated with immune infiltration (**Supplementary Table 11**), especially proportions of T-helper (Th2) cells (p-value = 2.8×10^−03^), dendritic cells (p-value = 1.8×10^−02^) and natural regulatory T (nTreg) cells (p-value = 2.3×10^−02^). Previous work established a connection between higher Th2 cells and progressive melanoma and in general are considered to be acting against immunotherapy by providing a microenvironment conducive to disease progression. Further, we found 8 out of the 29 identified proteins to be putative cancer driver genes (p-value = 4.1×10^−06^). Among 20 SKCM cell lines, we found 8 genes showing evidence for essentiality of cell proliferation (**Supplementary Table 12**), although there might be important etiological differences between BCC and SKCM.

## Discussion

In summary, we applied two complementary approaches, namely pQTS and ARCHIE, to identify the downstream plasma proteins with levels altered by the genetic variants associated with 21 different cancers. Results from applying pQTS identify several novel proteins through which the effect of multiple risk-variants are mediated for different cancers. ARCHIE provides further information on broader protein networks through which genetic associations may be mediated. Cross-cancer comparisons revealed shared proteins involved in the etiology of two or more cancers. Through extensive downstream analysis of two example cancers, cervical cancer and basal cell carcinoma, we show that the target proteins identified by both approaches are highly enriched in cancer related pathways, are enriched for expression in relevant cell populations, and different ARCHIE components can pinpoint distinct pathogenesis for the same cancer site. Additional evidence from data on somatic alternations, molecular profiling of related tumors and gene knockout experiments further validates the relevance of the identified target proteins.

The “omnigenic” model proposes that the effects of disease-related genetic variants converge on and is mediated via a smaller set of core genes^26,27^. Both our approaches, pQTS and ARCHIE are designed to capture such accumulation of effects of multiple cancer-related variants on plasma protein levels. In particular, pQTS aggregates the overall genetic risk of a cancer through the PRS and associates it to the protein levels. This can be conceptualized as a potential “retrospective” approach, where we find “optimal” predictor of the disease, via the PRS, and identify its association to plasma protein levels thereby potentially aggregating multiple pQTL associations. This is in contrast to the usual “prospective” approach adopted by TWAS or PWAS^8,10,68^, where we first identify the “optimal” predictor of the molecular phenotype (gene expression, proteins, and others) which is then tested for association with the disease. While the latter has a causal interpretation by following broad idea of “forward” genetics, it has been primarily restricted to analysis of cis effects of the SNPs. In contrast, pQTS can be applied genome-wide to identify both cis and trans associations and importantly, can be used to identify the convergence of multiple effects from the disease-related SNPs.

Recently Smith-Byrne et al^21^ used such a forward genetic approach to map potential causal proteins for 9 cancers based on cis-pQTLs associated with plasma proteins. Several of their findings were replicated including C4A, MICA and MICB associated with multiple cancers as well as ASIP with skin cancers (basal cell, squamous cell carcinoma and melanoma in our analysis). Overall, 18 of the 40 cancer related proteins identified in their analysis, were identified in either pQTS or ARCHIE in our analysis showing encouraging corroboration of evidence from independent analyses and distinct cohorts highlighting the potential of the proteins to be biologically relevant.

It is expected that in a complex disease like cancer the disease-associated genetic variants impact several different and distinct biological processes. PRS provides a reliable estimate of the genetic burden of the disease, and it can be used for powerful identification of the most downstream proteins through which effects of many variants are mediated. The pQTS approach, however, may miss more upstream genes in networks which may mediate effects of smaller groups of genetic variants. Thus, ARCHIE attempts to identify subsets of disease related SNPs that map to distinct protein (or gene) through trans-associations. The approach leads to natural partitioning of the SNPs constituting a PRS into subsets. Such partitioning of PRS have been previously conducted for partition type-2 diabetes PRS through analysis of glycemic biomarkers revealing distinct molecular mechanisms^69^. In the current analysis, the ARCHIE based partitioning of the BCC risk variants showcase the power of plasma proteomic data to partition PRS of cancer into clearly distinct etiological processes.

Our analysis using pQTS and ARCHIE replicates previously reported mechanisms, produces several novel results as well as improves interpretation of the PRS related to different cancers. Nevertheless, our analysis has several important limitations. First, our analysis may have modest power due to the limited sample size in our plasma protein reference (N=7,218 in self-reported White participants) as well as due to small effects associated with *trans*-associations. Both pQTS and ARCHIE, however, aggregates multiple *trans*-effects and thus have more power than standard single-SNP based *trans*-pQTL analyses. Further, plasma may fail to capture proteomic signatures from relevant tissue and cell types for many cancers, making the current findings potentially be biased toward certain proteins and processes, such as immunologic mechanisms which are better captured in plasma. In spite of these limitations, both pQTL based MR studies and direct association studies for cancer risk both have shown that plasma protein contains signatures of risk of many cancer sites, including those of solid cancers^16,21^. Nevertheless, it will be important to expand these analyses using multi-omics data from different tissue and cell types to obtain more comprehensive insight on biomarker networks through which genetic risks of cancers are modulated. Finally, while our analysis using the African American sample did provide evidence of replication for the discoveries made using the European American sample, the power was severely limited due to sample size and known transportability issues for current PRS for non-European ancestry populations.

The current research can lead to notable additions in biomarker development and identifying proteomic targets for cancers. Given that the proteome has proven to be suitable for drug targeting, identifying potential actionable targets would be crucial in follow up drug repurposing studies. Additionally, using the ARCHIE components to partition cancer-related SNPs into specific protein associated networks and can uncover the underlying heterogeneity captured by the disease or cancer related genetic variants. Further studies focus on exploring the gene-environment interactions of such partitioned SNP-sets to improve power over standard single-variant analysis while offering enhanced biological interpretation in terms of the corresponding protein network. However, the results need careful interpretation since variation in plasma protein levels can be attributable to a multitude of factors including differential binding affinity.

In summary, here we have used two approaches (pQTS and ARCHIE) to triangulate evidence for proteins associated with cancer related SNPs. Our methods capture different patterns of convergence of association signals on proteins. While pQTS identifies association(s) with the overall genetic burden, ARCHIE partitions the disease related SNPs into specific protein sets thereby improving the interpretation of the risk due to the SNPs. In general, this overall concept of convergence of association signals can be used to identify targets for any molecular phenotype. More importantly, the improved interpretation provided by identifying subclusters within the disease related SNPs, can produce novel insights into the distinct biological processes altered by the disease related SNPs. As newer omics datasets emerge, such complementary approaches can be useful to understand the overall genetic architecture of complex diseases as well as identify the downstream mechanisms of action of disease associated variants.

## Data Availability

All data produced are available online at dbGAP

## Data and resource availability

candidate cancer driver genes/proteins were curated from COSMIC (https://cancer.sanger.ac.uk/cosmic) and Network of Cancer Genes (http://network-cancer-genes.org/). CRISPR-screen data was collected from DepMap portal (https://depmap.org/portal/). scDRS (https://martinjzhang.github.io/scDRS/) was conducted on the basis of TMS data from <u>figshare</u>. Pathway analysis and additional bioinformatic analysis was conducted using online tools like ShinyGO: https://bioinformatics.sdstate.edu/go/ and GSCA: https://guolab.wchscu.cn/GSCA/.

## Funding and Acknowledgements

The ARIC study has been funded in whole or in part with Federal funds from the National Heart, Lung, and Blood Institute (NHLBI), National Institutes of Health (NIH), Department of Health and Human Services, under Contract nos. 75N92022D00001, 75N92022D00002, 75N92022D00003, 75N92022D00004, and 75N92022D00005. Studies on cancer in ARIC are supported by National Cancer Institute (NCI) grant U01 CA164975. This work was supported in part by NIH/NHLBI grant R01 HL134320. N.C. was supported by R01HG010480, U01 CA249866, 1R01HG013137. A.B was supported by R35GM139580 from National Institute of General Medical Sciences. D.D., M.J.M and K.B. were supported by the Intramural Research Program of the National Cancer Institute, National Institutes of Health. The authors thank the staff and participants of the ARIC study for their important contributions. SomaLogic Inc. conducted the SomaScan assays in exchange for use of ARIC data. Cancer data were provided by the Maryland Cancer Registry, Center for Cancer Prevention and Control, Maryland Department of Health, with funding from the state of Maryland and the Maryland Cigarette Restitution Fund. The collection and availability of cancer registry data are also supported by the Cooperative Agreement NU58DP007114, funded by the Centers for Disease Control and Prevention.

## Methods

### Study populations

The ARIC study is an ongoing community-based cohort study of individuals that initially enrolled 15,792 participants 1987 and 1989 from four communities across the US: Washington County, Maryland; suburbs of Minneapolis, Minnesota; Forsyth County, North Carolina; and Jackson, Mississippi. The second visit (v2) occurred in 1990–1992, when blood samples used for the measurement of the proteome were collected. The relative concentrations of plasma proteins or protein complexes from the blood samples were measured by SomaLogic Inc. using the version 4 platform by an aptamer (SOMAmer)-based approach. The details of available proteome data has been mentioned previously. Genotyping of ARIC samples was performed on the Affymetrix 6.0 DNA microarray and imputed to the TOPMed reference panel (Freeze 5b). We utilize available plasma proteome data measured on ARIC participants along with imputed genotypes, (N = 2,024 and 6,640 for AA and EA, respectively) to explore relationship between cancer PRS and plasma proteome.

### Cancer Ascertainment

Incident cancers in participants without a history of cancer at Visit 1 were identified by linking with state cancer registries in Maryland, Minnesota, Mississippi, and North Carolina, and by abstracting medical records and archived hospital discharge summaries, and death certificates^70^. In the analysis, we excluded first primary cancers diagnosed after Visit 2 through December 31, 2015. This resulted in 6,681 EA and 1,817 AA cancer-free participants for the final analysis.

### Preprocessing and QC

We used a previously developed QC pipeline for preprocessing, transforming and adjustment of known and hidden confounder effects for association analysis of the ARIC proteome data^10^. Briefly, genotyping of ARIC samples was performed on the Affymetrix 6.0 DNA microarray and imputed to the TOPMed reference panel (Freeze 5b). Further quality control metrics were implemented as detailed previously. The log-transformed relative abundances of SOMAmers were adjusted in a linear regression model including PEER factors (90 for EA and 80 for AA) and the covariates sex, age, study site and ten genetic PCs. The residuals from this linear regression were then rank-inverse normalized to avoid the influence of extreme values and were used as the corrected-protein quantification in the analysis.

### Construction of Cancer PRS

We analyzed 21 common cancers which have available PRS on PGScatalog (accessed on: September 2022) and have been associated with at least 10 independent common SNPs. Based on the prespecified weights downloaded from PGS catalog, we constructed PRS for each cancer for 9,084 individuals (7213 EA and 1871 AA) in ARIC v2.

### pQTS: association of PRS with protein levels

The PRS for diseases is a linear prediction model trained and tuned based on external GWAS. For a given cancer *C* the polygenic score for individual *i*, is given by:

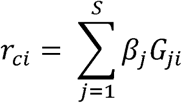

where *β_j_* is the weight or effect size of the *j^th^* SNP estimated from external GWAS, *G_ji_* is the genotype of the *j^th^* SNP for the *i^th^* individual (counted as the number of effect/risk alleles) and *s* is the number of SNPs constituting the PRS. The overall association of *r_ci_* with a protein level is given by the simple linear regression model:

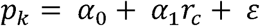

where *p_k_* is the normalized covariate adjusted level of the *k*^th^ protein for n individuals, *r_c_* - (*r*_*c*1_,*r*_*C*2_,…, *r_cn_*)’ is the vector of PRS for the same individuals, *ε* is a gaussian error; *α*_0_ and *α*_1_ are regression parameters. If the p-value for testing the null hypothesis *H*_0_: *α*_1_ - 0 vs *H*_0_: *α*_1_ ≠ 0 is significant, we denote the corresponding association as a pQTS. This test is performed across all proteins for a given cancer.

### Trans-association with PRS

For evaluating the trans-associations of the protein with the PRS of a given cancer, we create a sub-PRS, by restricting the PRS to SNPs which are physically distal (> 2.5Mb) from the transcription start site of the protein. We evaluate the association of this sub-PRS with the protein using the same linear model as above. This test is performed across all available proteins for a given cancer.

### Trans-association with ARCHIE

ARCHIE uses sparse canonical correlation analysis (sCCA) to identify subsets of disease-related SNPs trans-associated with selected sets of proteins. Given a set of SNPs constituting the PRS for a particular cancer, we look to identify sparse linear combinations of SNPs (*u*, termed SNP-component) which are strongly correlated sparse linear combinations of proteins (*v*, termed protein-component). In particular, we estimate the SNP and protein components (*u*, *v*; termed ARCHIE component together) using the following optimization:

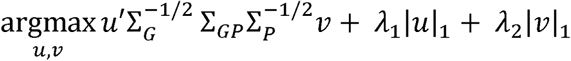

where Σ*_G_* is the LD matrix between the *s* SNPs, Σ*_P_* is the correlation matrix across the p proteins, Σ*_GP_* is the cross-correlation matrix between the *s* SNPs and *k* proteins, obtained from pairwise linear regression; *λ*_1_ and *λ*_2_ are sparsity parameters and |. |_1_ denote the *L_1_* regularization. At suitable levels of sparsity parameters, the SNPs (or proteins) corresponding to the non-zero entries of *u* (or *v*) are the SNPs (or proteins) selected by ARCHIE. We choose the sparsity parameters such that there is no overlap among the selected SNPs (or proteins) in successive components. We further define the squared canonical correlation (cc-value) captured by the components as

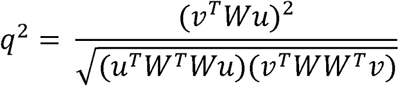

where 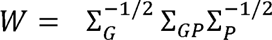. The cc-value is a measure of the overall trans-association captured by the SNPs and proteins selected in the ARCHIE component (*u*, *v*). The subsequent lower components of ARCHIE are obtained by deflating the matrix as *W_new_* - *W* - |*q*|*uv*’. The algorithm to estimate ARCHIE components (*u*,*v*) have been outlined previously.

### Competitive testing to identify significant ARCHIE components

To identify which ARCHIE components significantly capture the disease-specific *trans-*associated protein-sets, we evaluated the results from the original ARCHIE analysis against a *competitive* null hypothesis for each cancer. Since GWAS identified variants are expected to be enriched for *trans-*pQTLs in general, we test whether the cc-values (*q*^2^) obtained in the original analysis are higher than that obtained using the data from a random set of GWAS-identified variants and proteins of similar size, that do not reflect any disease-specific pattern. For this, we first construct a *null matrix* by taking a random sample of *s* variants from the pool of all variants available and extracting the corresponding *trans-*summary statistics for another set of randomly chosen *p* proteins. Then we use ARCHIE with the same sparsity levels as the original analysis, to extract the gene and variant components and calculate corresponding cc values. We repeat this step multiple (M) times to generate a competitive null distribution of cc values, allowing us to calculate a p-value for the observed cc-value in the original ARCHIE analysis for the disease under consideration.

### Tumor infiltration of Immune Cells

We used the data on tumor infiltration of immune cells reported by Liu et al for TCGA samples across different cancer types. The authors estimated the infiltrates of 24 immune cell types were evaluated through ImmuCellAI. The immune infiltration representing the proportion of a given immune cell type in a specific cancer type is then tested for correlation with the presence or absence of somatic mutation in at least one of the target genes (corresponding to the target proteins). We performed these analyses using the freely available GSCA pipeline.

### Cancer driver genes

We curated a list of 3356 canonical or candidate cancer driver genes from the Network of Cancer Genes & Healthy Drivers v7.0. Of these, 2986 had corresponding protein levels present in ARIC. Given the set of target proteins for a particular cancer, we use a Fisher’s exact test to evaluate whether the target proteins are overrepresented in the list of cancer driver genes.

### CRISPR essentiality screens

To investigate the effect of the genes underlying the target proteins on essentiality for proliferation and survival of cancer cells, we used the Achilles dataset from the DepMap portal. This provides estimated gene-dependency levels from CRISPR–Cas9 essentiality screens for a total 18,119 genes using a computational method, CERES. We first identified the genes among the targets identified by pQTS and/or ARCHIE for essentiality using a CERES value of −0.5 or lower as recommended. Further, we performed a bootstrapping experiment to determine the essentiality of the identified target genes. For a given cell line and cancer, we randomly chose the same number of target genes from the screens and calculated the average CERES score across all the genes. This was repeated multiple times to generate a distribution of average CERES expected from randomly selected genes. This allowed us to compute a p-value for the observed average CERES score for the identified target genes.

## Notes

### Competing Interest Statement

The authors have declared no competing interest.

## References

1. Buniello, A. et al. The NHGRI-EBI GWAS Catalog of published genome-wide association studies, targeted arrays and summary statistics 2019. Nucleic Acids Res 47, D1005–D1012 (2019).

2. Abdellaoui, A., Yengo, L., Verweij, K. J. H. & Visscher, P. M. 15 years of GWAS discovery: Realizing the promise. American Journal of Human Genetics vol. 110 Preprint at 10.1016/j.ajhg.2022.12.011 (2023).

3. Breen, G. et al. Translating genome-wide association findings into new therapeutics for psychiatry. Nature Neuroscience vol. 19 1392–1396 (2016).

4. Watanabe, K., Taskesen, E., van Bochoven, A. & Posthuma, D. Functional mapping and annotation of genetic associations with FUMA. Nat Commun 8, 1826 (2017).

5. Finucane, H. K. et al. Partitioning heritability by functional annotation using genome-wide association summary statistics. Nat Genet 47, 1228–1235 (2015).

6. Visscher, P. M. et al. 10 Years of GWAS Discovery: Biology, Function, and Translation. The American Journal of Human Genetics 101, 5–22 (2017).

7. Yin, X., et al. Integrating transcriptomics, metabolomics, and GWAS helps reveal molecular mechanisms for metabolite levels and disease risk. Am J Hum Genet 109 (10): 1727–1741 (2022).

8. Gusev, A. et al. Integrative approaches for large-scale transcriptome-wide association studies. Nat Genet 48, 245–252 (2016).

9. Wainberg, M. et al. Opportunities and challenges for transcriptome-wide association studies. Nat Genet 51 (4) 592–599 (2019).

10. Zhang, J. et al. Plasma proteome analyses in individuals of European and African ancestry identify cis-pQTLs and models for proteome-wide association studies. Nat Genet 54, (2022).

11. GTEx Consortium. The GTEx Consortium atlas of genetic regulatory effects across human tissues. Science (1979) 369, 1318–1330 (2020).

12. Sun, B. B. et al. Genomic atlas of the human plasma proteome. Nature 558, 73–79 (2018).

13. Sun, B. B. et al. Plasma proteomic associations with genetics and health in the UK Biobank. Nature 622, (2023).

14. The ARIC Investigators. The Atherosclerosis Risk in Communities (ARIC) Study: design and objectives. The ARIC investigators. Am J Epidemiol 129, 687–702 (1989).

15. Ferkingstad, E. et al. Large-scale integration of the plasma proteome with genetics and disease. Nat Genet 53, (2021).

16. Gadd, D. A. et al. Blood protein assessment of leading incident diseases and mortality in the UK Biobank. Nat Aging 4, 939–948 (2024).

17. Deng, Y.-T. et al. Atlas of the plasma proteome in health and disease in 53,026 adults. Cell 188, 253–271.e7 (2025).

18. Pietzner, M. et al. Mapping the proteo-genomic convergence of human diseases. Science (1979) 374, (2021).

19. Koprulu, M., et al. Proteogenomic links to human metabolic diseases. Nat Metab 5, (2023).

20. Zhao, J. H. et al. Genetics of circulating inflammatory proteins identifies drivers of immune-mediated disease risk and therapeutic targets. Nat Immunol 24, (2023).

21. Smith-Byrne, K. et al. Identifying therapeutic targets for cancer among 2074 circulating proteins and risk of nine cancers. Nat Commun 15, 3621 (2024).

22. Loesch, D. P. et al. Identification of plasma proteomic markers underlying polygenic risk of type 2 diabetes and related comorbidities. medRxiv 2024.03.15.24304200 (2024) doi:10.1101/2024.03.15.24304200.

23. Ritchie, S. C., et al. Integrative analysis of the plasma proteome and polygenic risk of cardiometabolic diseases. Nat Metab 3, (2021).

24. Yu, Z. et al. Polygenic risk scores for kidney function and their associations with circulating proteome, and incident kidney diseases. Journal of the American Society of Nephrology 32, (2021).

25. Võsa, U. et al. Large-scale cis- and trans-eQTL analyses identify thousands of genetic loci and polygenic scores that regulate blood gene expression. Nat Genet 53, 1300–1310 (2021).

26. Boyle, E. A., Li, Y. I. & Pritchard, J. K. An Expanded View of Complex Traits: From Polygenic to Omnigenic. Cell 169, 1177–1186 (2017).

27. Liu, X., Li, Y. I. & Pritchard, J. K. Trans Effects on Gene Expression Can Drive Omnigenic Inheritance. Cell 177, 1022–1034.e6 (2019).

28. Dutta, D. et al. Aggregative trans-eQTL analysis detects trait-specific target gene sets in whole blood. Nat Commun 13, 4323 (2022).

29. Lambert, S. A. et al. Enhancing the Polygenic Score Catalog with tools for score calculation and ancestry normalization. Nat Genet 56, 1989–1994 (2024).

30. Lambert, S. A. et al. The Polygenic Score Catalog as an open database for reproducibility and systematic evaluation. Nature Genetics vol. 53 Preprint at 10.1038/s41588-021-00783-5 (2021).

31. Hardoon, D. R. & Shawe-Taylor, J. Sparse canonical correlation analysis. Mach Learn 83, 331–353 (2011).

32. Witten, D. M. & Tibshirani, R. J. Extensions of Sparse Canonical Correlation Analysis with Applications to Genomic Data. Stat Appl Genet Mol Biol 8, 1–27 (2009).

33. Witten, D. M., Tibshirani, R. & Hastie, T. A penalized matrix decomposition, with applications to sparse principal components and canonical correlation analysis. Biostatistics 10, 515–534 (2009).

34. Sondka, Z. et al. The COSMIC Cancer Gene Census: describing genetic dysfunction across all human cancers. Nature Reviews Cancer vol. 18 Preprint at 10.1038/s41568-018-0060-1 (2018).

35. Repana, D. et al. The Network of Cancer Genes (NCG): A comprehensive catalogue of known and candidate cancer genes from cancer sequencing screens. Genome Biol 20, (2019).

36. Li, Q. & Lozano, G. Molecular pathways: Targeting Mdm2 and Mdm4 in cancer therapy. Clinical Cancer Research 19, (2013).

37. Danovi, D. et al. Amplification of Mdmx (or Mdm4) Directly Contributes to Tumor Formation by Inhibiting p53 Tumor Suppressor Activity. Mol Cell Biol 24, (2004).

38. Liu, J. et al. MDM4 was associated with poor prognosis and tumor-immune infiltration of cancers. Eur J Med Res 29, (2024).

39. Wang, H. et al. The Proto-oncogene c-Kit Inhibits Tumor Growth by Behaving as a Dependence Receptor. Mol Cell 72, (2018).

40. Shi, X. et al. Distinct cellular properties of oncogenic KIT receptor tyrosine kinase mutants enable alternative courses of cancer cell inhibition. Proc Natl Acad Sci U S A 113, (2016).

41. Huang, J. et al. Cytokine-like molecule CCDC134 contributes to CD8+ T-cell effector functions in cancer immunotherapy. Cancer Res 74, (2014).

42. Jamieson, T. et al. Inhibition of CXCR2 profoundly suppresses inflammation-driven and spontaneous tumorigenesis. Journal of Clinical Investigation 122, (2012).

43. Saintigny, P. et al. CXCR2 expression in tumor cells is a poor prognostic factor and promotes invasion and metastasis in lung adenocarcinoma. Cancer Res 73, (2013).

44. Li, Y. et al. Targeting IL-21 to tumor-reactive T cells enhances memory T cell responses and anti-PD-1 antibody therapy. Nat Commun 12, (2021).

45. Rees, M. G. et al. Correlating chemical sensitivity and basal gene expression reveals mechanism of action. Nat Chem Biol 12, (2016).

46. Sirugo, G., Williams, S. M. & Tishkoff, S. A. The Missing Diversity in Human Genetic Studies. Cell vol. 177 Preprint at 10.1016/j.cell.2019.02.048 (2019).

47. Martin, A. R. et al. Clinical use of current polygenic risk scores may exacerbate health disparities. Nat Genet 51, (2019).

48. Bowden, S. J. et al. Genetic variation in cervical preinvasive and invasive disease: a genome-wide association study. Lancet Oncol 22, (2021).

49. Madeleine, M. M. et al. Human leukocyte antigen class II and cervical cancer risk: A population-based study. Journal of Infectious Diseases 186, (2002).

50. Adebamowo, S. N., et al. Genome, HLA and polygenic risk score analyses for prevalent and persistent cervical human papillomavirus (HPV) infections. European Journal of Human Genetics (2024) doi:10.1038/s41431-023-01521-7.

51. Graff, R. E. et al. Cross-cancer evaluation of polygenic risk scores for 16 cancer types in two large cohorts. Nat Commun 12, 970 (2021).

52. Yan, S. P. et al. LncRNA LINC01305 silencing inhibits cell epithelial-mesenchymal transition in cervical cancer by inhibiting TNXB-mediated PI3K/Akt signalling pathway. Journal of Cellular and Molecular Medicine vol. 23 Preprint at 10.1111/jcmm.14161 (2019).

53. Chen, J. R. et al. MHC class I chain-related gene a (MICA) polymorphism and the different histological types of cervical cancer. Neoplasma 52, (2005).

54. Pollock, N. R., Harrison, G. F. & Norman, P. J. Immunogenomics of Killer Cell Immunoglobulin-Like Receptor (KIR) and HLA Class I: Coevolution and Consequences for Human Health. Journal of Allergy and Clinical Immunology: In Practice 10, (2022).

55. Khakoo, S. I. & Jamil, K. M. KIR/HLA interactions and pathogen immunity. Journal of Biomedicine and Biotechnology vol. 2011 Preprint at 10.1155/2011/298348 (2011).

56. Bao, X. et al. HLA and KIR associations of cervical neoplasia. Journal of Infectious Diseases 218, (2018).

57. Wilson, T. J. et al. FcRL6, a new ITIM-bearing receptor on cytolytic cells, is broadly expressed by lymphocytes following HIV-1 infection. Blood 109, (2007).

58. Schreeder, D. M. et al. Cutting Edge: FcR-Like 6 Is an MHC Class II Receptor. The Journal of Immunology 185, (2010).

59. Zhang, M. et al. Identification of NEO1 as a prognostic biomarker and its effects on the progression of colorectal cancer. Cancer Cell Int 20, (2020).

60. Yuxiong, W. et al. Regulatory mechanisms of the cAMP-responsive element binding protein 3 (CREB3) family in cancers. Biomedicine and Pharmacotherapy vol. 166 Preprint at 10.1016/j.biopha.2023.115335 (2023).

61. Pu, Q. et al. The Novel Transcription Factor CREB3L4 Contributes to the Progression of Human Breast Carcinoma. J Mammary Gland Biol Neoplasia 25, (2020).

62. Kim, T.-H., Park, J.-M., Kim, M.-Y. & Ahn, Y.-H. The role of CREB3L4 in the proliferation of prostate cancer cells. Sci Rep 7, 45300 (2017).

63. Zhang, M. J. et al. Polygenic enrichment distinguishes disease associations of individual cells in single-cell RNA-seq data. Nat Genet 54, (2022).

64. Weinstein, J. N. et al. The cancer genome atlas pan-cancer analysis project. Nat Genet 45, (2013).

65. Chahal, H. S. et al. Genome-wide association study identifies 14 novel risk alleles associated with basal cell carcinoma. Nat Commun 7, (2016).

66. Zhang, S. et al. PMEL as a Prognostic Biomarker and Negatively Associated with Immune Infiltration in Skin Cutaneous Melanoma (SKCM). Journal of Immunotherapy 44, (2021).

67. Choquet, H., et al. Multi-ancestry genome-wide meta-analysis identifies novel basal cell carcinoma loci and shared genetic effects with squamous cell carcinoma. Commun Biol 7, (2024).

68. Gamazon, E. R. et al. A gene-based association method for mapping traits using reference transcriptome data. Nat Genet 47(9), 1091–1098 (2015).

69. Dicorpo, D. et al. Type 2 Diabetes Partitioned Polygenic Scores Associate With Disease Outcomes in 454,193 Individuals Across 13 Cohorts. Diabetes Care 45, (2022).

70. Joshu, C. E. et al. Enhancing the infrastructure of the atherosclerosis risk in Communities (ARIC) study for cancer epidemiology research: Aric cancer. Cancer Epidemiology Biomarkers and Prevention 27, (2018).

